# RVOT Reconstruction in d-Transposition of the Great Arteries with Ventricular Septal Defect and Pulmonary Obstruction: Male-Female Differences in Clinical and Homograft Function

**DOI:** 10.1101/2023.10.18.23297234

**Authors:** Xu Wang, Isabelle M. Bennink, Kevin M. Veen, Pieter C. van de Woestijne, Carlijn C. E. M. van der Ven, Ad J. J. C. Bogers, Eleni-Rosalina Andrinopoulou, Wim. A. Helbing, Johanna J.M. Takkenberg

## Abstract

**Background:** This study investigated male-female differences in clinical and homograft outcomes in dextro-transposition of the great arteries (d-TGA) patients, with ventricular septal defect (VSD) and pulmonary stenosis/atresia (PS/PA), who underwent right ventricular outflow tract (RVOT) reconstruction with a homograft.

**Methods:** All d-TGA with VSD and PS/PA patients receiving a homograft for RVOT reconstruction at our center from 1986 to 2021 were included. Time-to-event analyses were used for time-related clinical outcomes and mixed-effects models to characterize homograft function.

**Results:** Twenty-four patients (16 males) in whom 38 homografts were implanted over time were included. Median age at the first homograft implantation was 2.56 years [IQR: 1.26-11.31] and 2.06 years [IQR: 1.28-8.13] for males and females. Two early death (both males) and five late deaths (2 males) were observed. Twenty-two RVOT reinterventions (19/3 in males/females) occurred in follow-up (males/females:345.72/173.35 patient-years). The 10-year survival probability was 87.5% (95% CI: 67.3-100) for females and 87.1% (95% CI: 71.8-100) for males. Freedom from RVOT reintervention at 10-year was 87.5% (95% CI: 67.3-100) for females and 64.9% (95% CI: 47.9-88.0) for males. RVOT peak gradient increase rate was associated with body growth rate (7.27 [95% CI: 2.08-12.45], P=0.0133).

**Conclusions:** Substantial male-female differences in clinical and homograft outcomes were found in d-TGA with VSD and PS/PA patients receiving a homograft for RVOT reconstruction. Male patients had a higher RVOT reintervention rate and faster progression to homograft stenotic failure compared to females. Oversized homografts might benefit children for the strong association between rates of homograft stenosis and body growth.

**Clinical Perspective:**

1. What is new?

- Male patients diagnosed with d-TGA, VSD, and PS/PA demonstrated a significantly higher rate of RVOT homograft reinterventions and a more rapid progression toward homograft stenotic failure when compared with their female counterparts.
- The male-female differences in clinical outcomes and homograft function may be attributed, in part, to the faster body growth observed in male children, especially given the strong association between rates of homograft stenosis and body growth.
2. What are the clinical implications?

- For rapidly growing young children, opting for a suitably oversized homograft might serve as a more effective solution in reducing the rate of homograft stenotic failure.
- By exploring the relationship between homograft stenosis rate and individual body growth rate for each child, a personalized approach can be adopted during the homograft selection process, optimizing the sizing strategy for a better clinical outcome.

## INTRODUCTION

Dextro-transposition of the great arteries(d-TGA) with ventricular septal defect (VSD) and pulmonary stenosis (PS) or pulmonary atresia (PA) is a rare congenital heart defect (CHD) that requires early palliative or corrective operations to enable patients’ survival^1^. Despite various corrective options available, dysfunction of the right ventricular outflow tract (RVOT) conduit is almost inevitable during a patient’s lifetime^2^. If feasible, implantation of a valve substitute with a homograft is the preferred choice to address this issue^3^. As most patients received their first homograft as a RVOT conduit during childhood, a period of rapid body growth and development, the homograft function evolution differs from that observed in adults^4^. Additionally, considering the distinct growth patterns between boys and girls^5^, there may be male-female differences in clinical outcomes and homograft function in d-TGA, VSD, PS/PA patients who received homograft as a RVOT conduit during childhood.

Despite the importance of studying this patient group, limited research has been done on these patients, particularly in terms of sex-specific outcomes. Thus, this study aimed to explore male-female differences in clinical outcomes and homograft function after homograft RVOT reconstruction in patients diagnosed with d-TGA, VSD, PS/PA.

## PATIENTS AND METHODS

This is a retrospective single-center study. The medical ethics committee (METC) of the Erasmus University Medical Center reviewed and approved this study (MEC-2012-477). Informed consent was obtained from all participants alive. No consent from deceased patients was necessary according to the medical ethics regulations of our medical center.

### 1. Study population

All d-TGA patients who underwent RVOT reconstruction with a homograft at Erasmus University Medical Center from April 1986 to December 2021 were identified. Patients not surgically corrected, without VSD and PS/PA, or alive but not giving their consents were excluded. Patients who underwent univentricular repair were also excluded.

### 2. Data collection

Baseline characteristics, perioperative information, clinical outcomes and echocardiographic parameters during follow-up were extracted by reviewing medical records. Events were defined according to the Akins 2008 guidelines ^6^. Early events were defined as all events occurring within the first 30 days after operation. Any events occurring after 30 days postoperative were defined as late events. Structural valve deterioration (SVD) was defined as stenosis or regurgitation relevant to pulmonary valve without diagnosis of endocarditis. The severities of stenosis and regurgitation were classified in compliance with the ESC/EACTS 2021 guidelines^7^. Stenosis was graded as mild (<36mmHg), moderate (36-64mmHg), and severe (>64mmHg). Regurgitation was graded as zero/trace, mild, moderate, and severe.

### 3. Statistical methods

Continuous data are presented as mean ± standard deviation (normal distribution) or median with interquartile range (IQR) (non-normal distribution). Categorical data are presented as absolute count with percentages. Comparisons among continuous variables were made with the independent samples t-test or Kruskal-Wallis test, as appropriate. Comparisons of categorical data were made with the Chi-square test or Fisher’s exact test, as appropriate^8^. The Shapiro-Wilk test and visual inspections were used to test the normality. The adverse event rate (AER) for clinical outcomes was calculated as total number of observed events over time (including repeated events) divided by total patient-years of follow-up. Survival data are presented as Kaplan-Meier estimates with a standard error or in the form of Kaplan-Meier curves. The log-rank test was used to compare survival outcomes in male and female patients. Fine-Gray sub-distribution hazard models were used for competing risk analysis, with death being the competing event, and the sex-specific cumulative incidence of homograft reintervention and endocarditis was plotted based on the results^9^.

Two echocardiographic parameters were analyzed to assess homograft function, with only sex, time, and their interaction term as covariates to prevent overfitting. The RVOT peak gradient was evaluated using linear mixed-effects models with random intercepts for homografts. The degree of regurgitation was dichotomized as < moderate or ≥moderate and analyzed using logistic mixed-effects models with random intercepts for homografts. In patients younger than 16 years old, the longitudinal evolution of RVOT peak gradient was correlated to the longitudinal evolution of body surface area (BSA) by multivariate (multiple outcomes) mixed modeling^10^. The correlation between RVOT peak gradient and BSA was taken into account via the random effects of the model, in particular a multivariate normal distribution was assumed for the variance-covariance matrix of the random effects. Additionally, random slopes of BSA and RVOT peak gradient were predicted in their respective linear mixed-effects models, and associations between them were explored in linear regression model. Sensitivity analysis was conducted by temporarily excluding all measurements of one homograft at a time (leave-one-out sensitivity analysis). The estimates and their 95% CIs were visualized respectively for each model parameter. Linear mixed-effects models with the same structure were then constructed, and the results were compared with the full model without any exclusion. A P-value less than 0.05 was considered statistically significant, and no correction for multiple testing was applied. Statistical analyses were performed with R statistical program (R Foundation for Statistical Computing, Version 4.2.2. https://www.R-project.org/).

## RESULTS

### 1. Baseline characteristics and perioperative information

A total of 24 patients (16 males, 8 females), in whom 38 homografts (29 in males, 9 in females) were used for RVOT reconstructions, were included. Patient baseline characteristics are presented in **Table 1**. The perioperative information for each homograft implantation procedures is provided in Supplementary material Table S1. Two patients died during hospitalizations (early death) for homograft replacement. Both of them were males: one died of refractory bleeding and hemorrhagic shock 3 days after operation; one died of septic shock 53 days after the operation. No other early endocarditis or reintervention for homograft was observed.

**Table 1:**
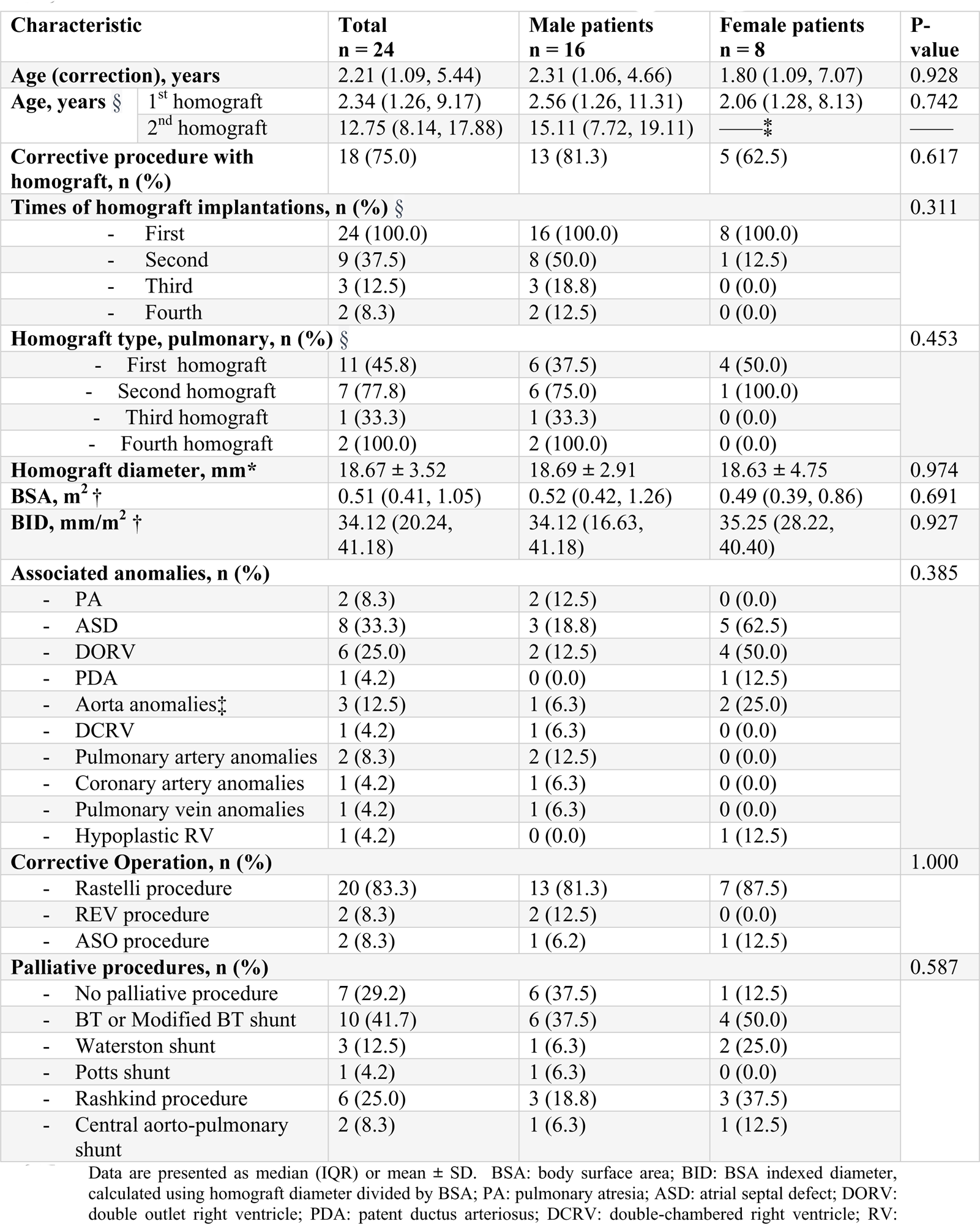
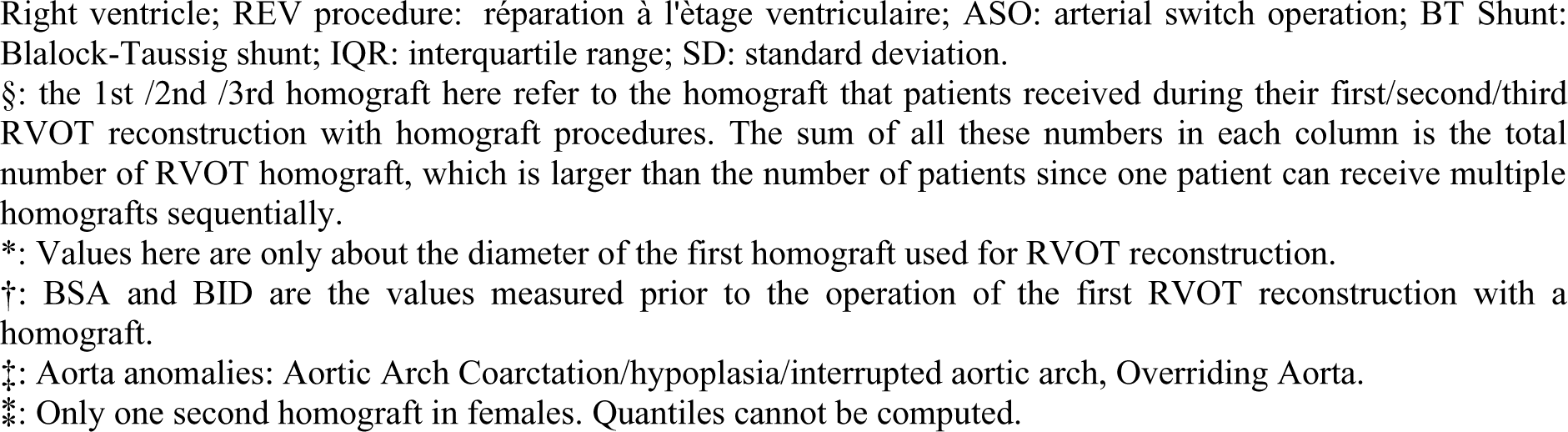
Baseline characteristics of d-TGA patients with VSD, pulmonary stenosis or atresia.

| Characteristic |  | Total<br>n = 24 | Male patients<br>n = 16 | Female patients<br>n = 8 | P-value |
| --- | --- | --- | --- | --- | --- |
| **Age (correction), years** |  | 2.21 (1.09, 5.44) | 2.31 (1.06, 4.66) | 1.80 (1.09, 7.07) | 0.928 |
| **Age, years §** | 1<sup>st</sup> homograft | 2.34 (1.26, 9.17) | 2.56 (1.26, 11.31) | 2.06 (1.28, 8.13) | 0.742 |
|  | 2<sup>nd</sup> homograft | 12.75 (8.14, 17.88) | 15.11 (7.72, 19.11) | ——* | —— |
| **Corrective procedure with homograft, n (%)** |  | 18 (75.0) | 13 (81.3) | 5 (62.5) | 0.617 |
| **Times of homograft implantations, n (%) §** |  |  |  |  | 0.311 |
| - First |  | 24 (100.0) | 16 (100.0) | 8 (100.0) |  |
| - Second |  | 9 (37.5) | 8 (50.0) | 1 (12.5) |  |
| - Third |  | 3 (12.5) | 3 (18.8) | 0 (0.0) |  |
| - Fourth |  | 2 (8.3) | 2 (12.5) | 0 (0.0) |  |
| **Homograft type, pulmonary, n (%) §** |  |  |  |  | 0.453 |
| - First homograft |  | 11 (45.8) | 6 (37.5) | 4 (50.0) |  |
| - Second homograft |  | 7 (77.8) | 6 (75.0) | 1 (100.0) |  |
| - Third homograft |  | 1 (33.3) | 1 (33.3) | 0 (0.0) |  |
| - Fourth homograft |  | 2 (100.0) | 2 (100.0) | 0 (0.0) |  |
| **Homograft diameter, mm\*** |  | 18.67 ± 3.52 | 18.69 ± 2.91 | 18.63 ± 4.75 | 0.974 |
| **BSA, m<sup>2</sup> †** |  | 0.51 (0.41, 1.05) | 0.52 (0.42, 1.26) | 0.49 (0.39, 0.86) | 0.691 |
| **BID, mm/m<sup>2</sup> †** |  | 34.12 (20.24, 41.18) | 34.12 (16.63, 41.18) | 35.25 (28.22, 40.40) | 0.927 |
| **Associated anomalies, n (%)** |  |  |  |  | 0.385 |
| - PA |  | 2 (8.3) | 2 (12.5) | 0 (0.0) |  |
| - ASD |  | 8 (33.3) | 3 (18.8) | 5 (62.5) |  |
| - DORV |  | 6 (25.0) | 2 (12.5) | 4 (50.0) |  |
| - PDA |  | 1 (4.2) | 0 (0.0) | 1 (12.5) |  |
| - Aorta anomalies‡ |  | 3 (12.5) | 1 (6.3) | 2 (25.0) |  |
| - DCRV |  | 1 (4.2) | 1 (6.3) | 0 (0.0) |  |
| - Pulmonary artery anomalies |  | 2 (8.3) | 2 (12.5) | 0 (0.0) |  |
| - Coronary artery anomalies |  | 1 (4.2) | 1 (6.3) | 0 (0.0) |  |
| - Pulmonary vein anomalies |  | 1 (4.2) | 1 (6.3) | 0 (0.0) |  |
| - Hypoplastic RV |  | 1 (4.2) | 0 (0.0) | 1 (12.5) |  |
| **Corrective Operation, n (%)** |  |  |  |  | 1.000 |
| - Rastelli procedure |  | 20 (83.3) | 13 (81.3) | 7 (87.5) |  |
| - REV procedure |  | 2 (8.3) | 2 (12.5) | 0 (0.0) |  |
| - ASO procedure |  | 2 (8.3) | 1 (6.2) | 1 (12.5) |  |
| **Palliative procedures, n (%)** |  |  |  |  | 0.587 |
| - No palliative procedure |  | 7 (29.2) | 6 (37.5) | 1 (12.5) |  |
| - BT or Modified BT shunt |  | 10 (41.7) | 6 (37.5) | 4 (50.0) |  |
| - Waterston shunt |  | 3 (12.5) | 1 (6.3) | 2 (25.0) |  |
| - Potts shunt |  | 1 (4.2) | 1 (6.3) | 0 (0.0) |  |
| - Rashkind procedure |  | 6 (25.0) | 3 (18.8) | 3 (37.5) |  |
| - Central aorto-pulmonary shunt |  | 2 (8.3) | 1 (6.3) | 1 (12.5) |  |

Data are presented as median (IQR) or mean ± SD. BSA: body surface area; BID: BSA indexed diameter, calculated using homograft diameter divided by BSA; PA: pulmonary atresia; ASD: atrial septal defect; DORV: double outlet right ventricle; PDA: patent ductus arteriosus; DCRV: double-chambered right ventricle; RV:
Right ventricle; REV procedure: réparation à l'étage ventriculaire; ASO: arterial switch operation; BT Shunt: Blalock-Taussig shunt; IQR: interquartile range; SD: standard deviation. §: the 1st /2nd /3rd homograft here refer to the homograft that patients received during their first/second/third RVOT reconstruction with homograft procedures. The sum of all these numbers in each column is the total number of RVOT homograft, which is larger than the number of patients since one patient can receive multiple homografts sequentially. \*: Values here are only about the diameter of the first homograft used for RVOT reconstruction. †: BSA and BID are the values measured prior to the operation of the first RVOT reconstruction with a homograft. ‡: Aorta anomalies: Aortic Arch Coarctation/hypoplasia/interrupted aortic arch, Overriding Aorta. \*: Only one second homograft in females. Quantiles cannot be computed.

### 2. Clinical outcomes

The median length of follow-up was 28.71 years (IQR: 7.30-31.57; patient-years: 345.72) in male patients and 26.24 years (IQR: 12.22-31.95; patient-years: 173.35) in female patients. The completeness of follow-up was 95.98 %, calculated by the method proposed by Wu et al.^11^. **Figure 1** displays the time slots of key d-TGA-related events in each patient’s journey from birth to death or the end of follow-up, separated by sex. These events include d-TGA corrective operation, first RVOT reconstruction with homograft, subsequent RVOT relevant procedures, homograft endocarditis, and mortality or end of follow-up.

**Figure 1:**
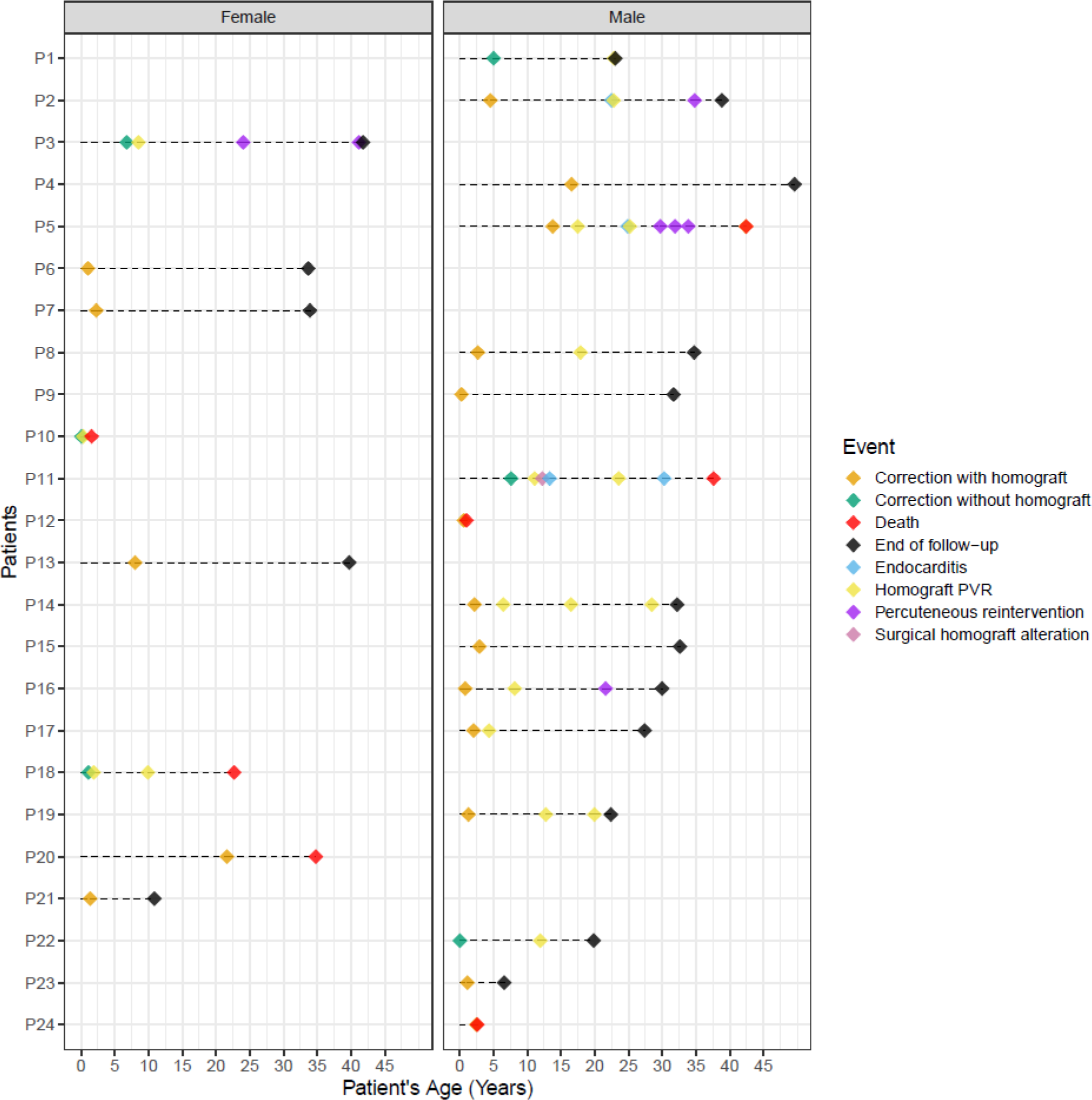
Event timelines of patients with d-TGA/VSD and PS or PA. Each point on the x-axis represents one patient. Each horizontal line stands for the time span of on patient from his or her birth to death or the end of the follow-up. Diamonds of different colors represent different events. All events were arranged chronologically, with the corresponding values on y-axis indicating the age of event occurrence. Patient “P5” died 3 days after the last homograft PVR operation. “PVR”: pulmonary valve replacement; “Homograft PVR” is a synonym for RVOT reconstruction with homograft.

#### 2.1 Late mortality

In total, five late deaths (2 males/3 females) occurred during follow-up. Reasons for three deaths in females were as follows: death while awaiting surgery for severe homograft stenosis, erysipelas-induced septic shock, and severe arrhythmia, respectively. Two male patients were found dead at home without clear explanations. The AERs of late death are presented in **Table 2**. The Kaplan-Meier estimates of overall survival are shown in **Figure 2**. The 5-,10-,15-, and 20-year survival probabilities of male and female patients are presented in **Table 3**.

**Figure 2:**
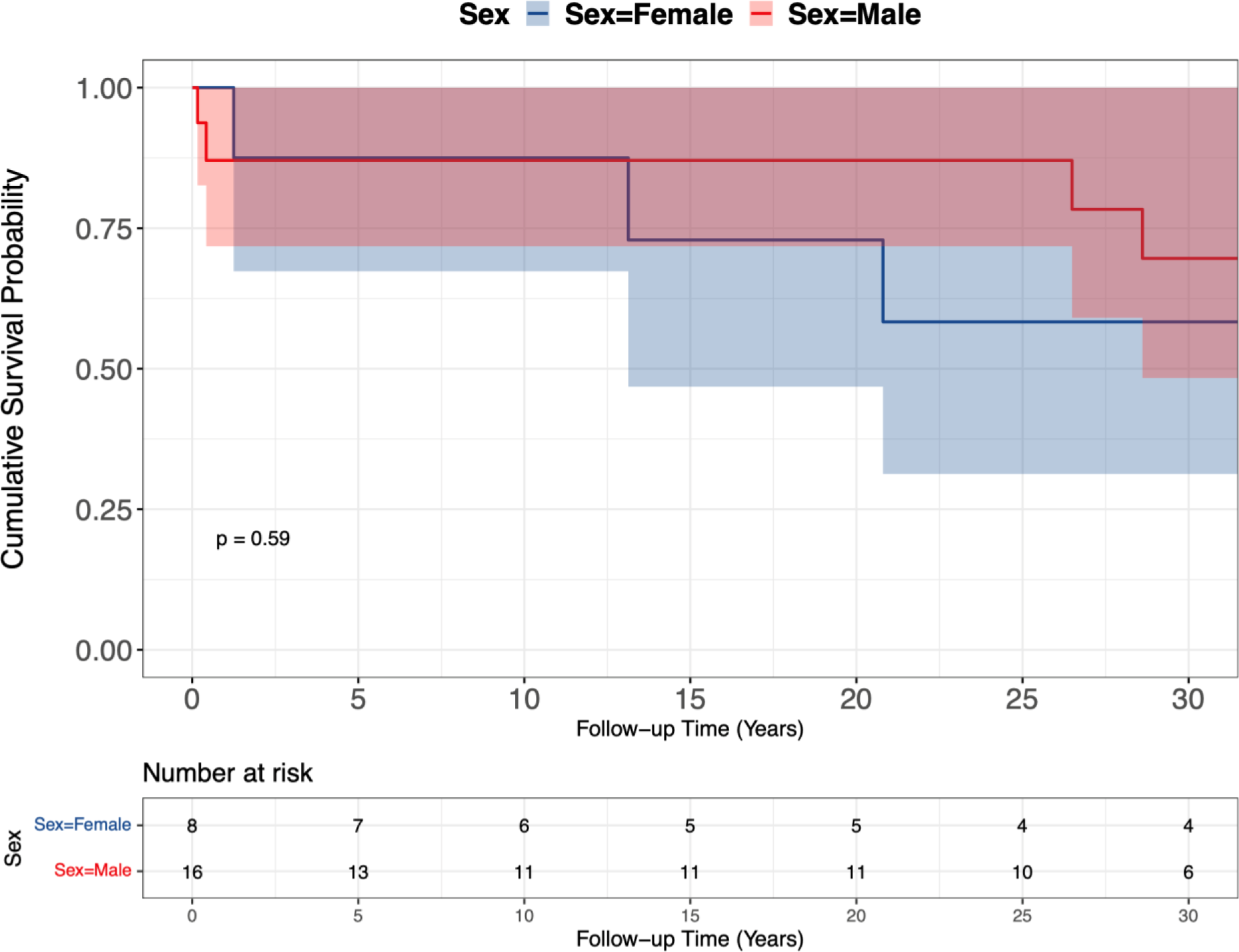
Kaplan-Meier survival estimates in male and female patients. The number-at-risk is the number of patients at risk at different time points(not the number of homograft). Log-rank test was used here.

**Table 2:** Number of events and adverse event rates in male and female patients.

|  |  | Males (n = 16) |  | Females (n = 8) |  | P-values |
| --- | --- | --- | --- | --- | --- | --- |
|  |  | N | AER (95%CI) | N | AER (95%CI) |  |
| FUP, years |  | 28.71 (7.30, 31.57) |  | 26.24 (12.22, 31.95) |  |  |
| Patient-years |  | 345.72 |  | 173.35 |  |  |
| Late mortality | All-cause | 2 | 0.58 (0.07-2.09) | 3 | 1.73 (0.36-5.06) | 0.207 |
|  | -Cardiac | 2 | 0.58 (0.07-2.09) | 2 | 1.15 (0.14-4.17) | 0.481 |
|  | -Valve-related | 2 | 0.58 (0.07-2.09) | 1 | 0.58 (0.01-3.21) | 0.998 |
| Late reinterventions |  | 19 | 5.50 (3.31-8.58) | 3 | 1.73 (0.36-5.06) | 0.049 |
| - Percutaneous |  | 5 | 1.45 (0.47-3.38) | 2 | 1.15 (0.14-4.17) | 0.787 |
| - Surgical |  | 14 | 4.05 (2.21-6.79) | 1 | 0.58 (0.01-3.21) | 0.028 |
| Replacement |  | 16 | 4.63 (2.65-7.52) | 1 | 0.58 (0.01-3.21) | 0.016 |
| - Percutaneous |  | 3 | 0.87 (0.18-2.54) | 0 | 0 (0-2.13) | 0.220 |
| - Surgical |  | 13 | 3.76 (2.00-6.43) | 1 | 0.58 (0.01-3.21) | 0.037 |
| Endocarditis |  | 4 | 1.16 (0.32-2.97) | 0 | 0 (0-2.13) | 0.157 |
| PPMI/ICD |  | 2 | 0.58 (0.07-2.09) | 1 | 0.58 (0.01-3.21) | 0.997 |

N: number of events; AER: adverse event rate, patient-years divided by number of events; CI: confidence interval; FUP: length of follow-up; PPMI: permean pacemaker implantation; ICD: Implantable cardioverter defibrillator.

**Table 3:** Probabilities of cumulative survival, and freedom from homograft reintervention, replacement and endocarditis.

| Event | Sex | 5 years | 10 years | 15 years | 20 years |
| --- | --- | --- | --- | --- | --- |
| **Patients Survival\***, % | Female | 87.5 (67.3-100) | 87.5 (67.3-100) | 72.9 (46.8-100) | 72.9 (46.8-100) |
|  | Male | 87.1 (71.8-100) | 87.1 (71.8-100) | 87.1 (71.8-100) | 87.1 (71.8-100) |
| **Freedom from reintervention†**, % | Female | 100 (100-100) | 87.5 (67.3-100) | 87.5 (67.3-100) | 66.5 (35.2-100) |
|  | Male | 83.4 (69.8-99.7) | 64.9 (47.9-88.0) | 38.1 (21.7-66.9) | 23.8 (9.9-57.1) |
| - *Percutaneous* | Female | 100 (100-100) | 100 (100-100) | 100 (100-100) | 75.0 (42.6-100) |
|  | Male | 100 (100-100) | 94.7 (85.2-100) | 74.6 (52.6-100) | 74.6 (52.6-100) |
| - *Surgical* | Female | 100 (100-100) | 87.5 (67.3-100) | 87.5 (67.3-100) | 87.5 (67.3-100) |
|  | Male | 83.4 (69.8-100) | 69.5 (52.9-91.3) | 53.8 (36.1-80.2) | 27.7(11.5-66.3) |
| **Freedom from replacement†**, % | Female | 100 (100-100) | 87.5 (67.3-100) | 87.5 (67.3-100) | 87.5 (67.3-100) |
|  | Male | 87.5 (75.1-100) | 69.0 (52.3-91.1) | 47.5 (29.9-75.5) | 29.7 (13.1-67.0) |
| - *Percutaneous* | Female | 100 (100-100) | 100 (100-100) | 100 (100-100) | 100 (100-100) |
|  | Male | 100 (100-100) | 94.7 (85.2-100) | 85.3 (67.6-100) | 85.3 (67.6-100) |
| - *Surgical* | Female | 100 (100-100) | 87.5 (67.3-100) | 87.5 (67.3-100) | 87.5 (67.3-100) |
|  | Male | 87.5 (75.1-100) | 73.6 (57.5-94.3) | 52.3 (34.4-79.6) | 26.9 (11.1-65.1) |
| **Freedom from endocarditis†**, % | Female | 100 (100-100) | 100 (100-100) | 100 (100-100) | 100 (100-100) |
|  | Male | 96.0 (88.6-100) | 85.3(71.0-100) | 85.3(71.0-100) | 71.1 (47.5-100) |

\*: Survival probability was calculated on patient-level. †: Probabilities of freedom from reintervention, replacement, and endocarditis were calculated on homograft-level. All probabilities are presented in the format of absolute percent (95% confidence interval).

#### 2.2 Homograft reintervention

During follow-up, 10 patients (8 males/2 females) received 22 reinterventions (7 percutaneous, 15 surgical). Of all reinterventions, 17 were pulmonary valve replacement (3 percutaneous, 14 surgical). The AERs of late reinterventions and replacement male and female patients are presented in **Table 2**. Most reinterventions were for stenotic SVD, and two homografts were replaced due to endocarditis. Most surgical replacement procedures were performed in male patients (13/14, 92.9%). Details of all reintervention cases are presented in Supplementary material Table S2.

The cumulative incidence of RVOT reintervention with death as a competing event was visualized and is presented in **Figure 3**. Male patients had a significant higher cumulative incidence of RVOT reintervention compared to female patients (Gray’s test: P=0.0296). The Kaplan-Meier estimates of freedom from RVOT replacement are shown in Supplementary material Figure S1. The 5-,10-,15-, and 20-year probabilities of freedom from RVOT reintervention and replacement in male and female patients are presented in **Table 3**. Additionally, the Kaplan-Meier estimates of freedom from RVOT percutaneous/surgical reintervention and percutaneous/surgical replacement are shown in Supplementary material Figure S2-S5.

**Figure 3:**
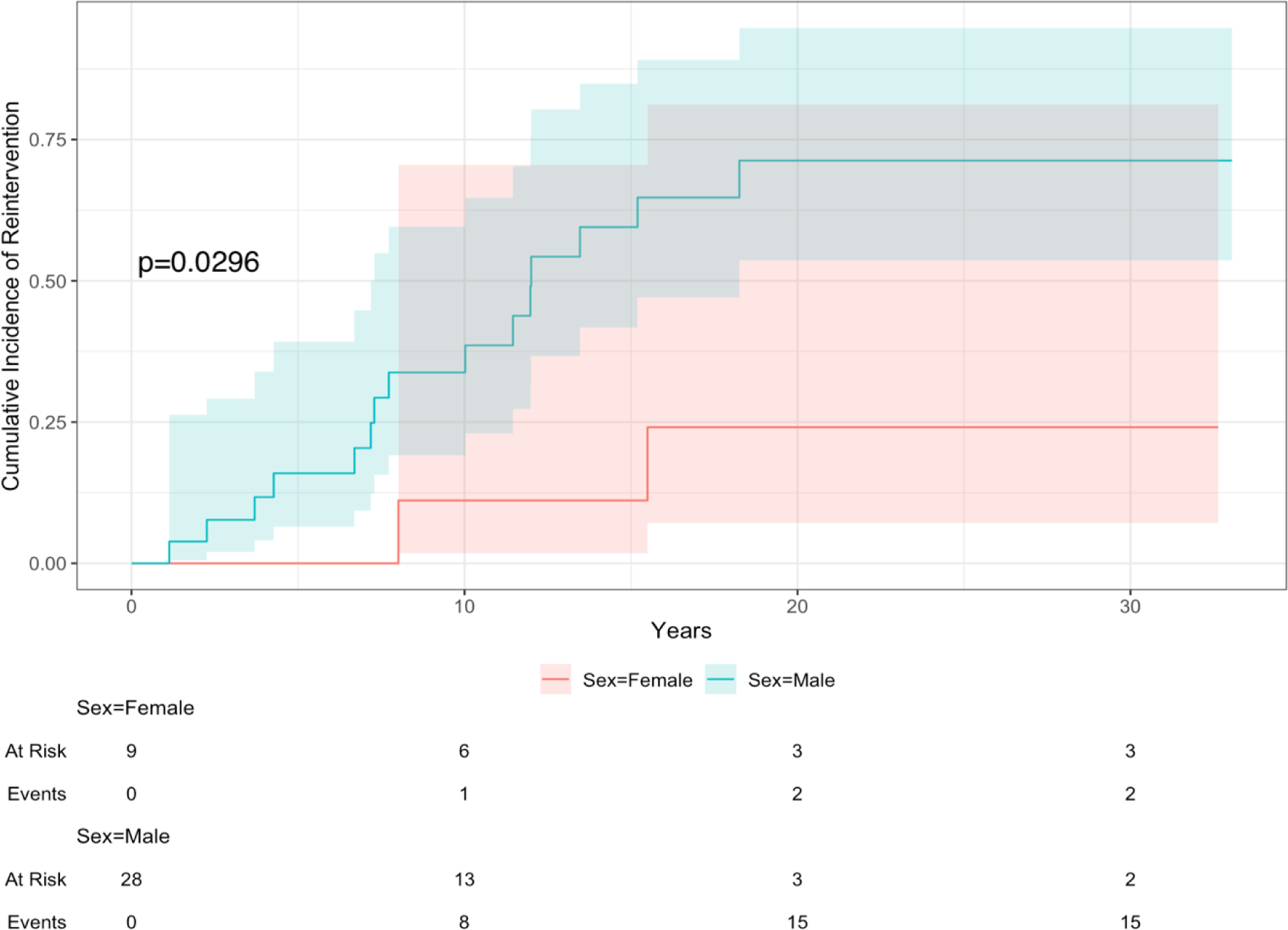
Cumulative incidence of receiving RVOT reinterventions in homografts implanted in male and female patients.

#### 2.3 Endocarditis

There were 4 homografts with endocarditis (all males), of which two required surgical homograft replacement. The AERs of endocarditis are presented in **Table 2**. The cumulative incidence of homograft endocarditis with death as the competing event was visualized and is presented in **Figure 4**. The 5-,10-,15-, and 20-year probabilities of freedom from endocarditis are presented in **Table 3**.

**Figure 4:**
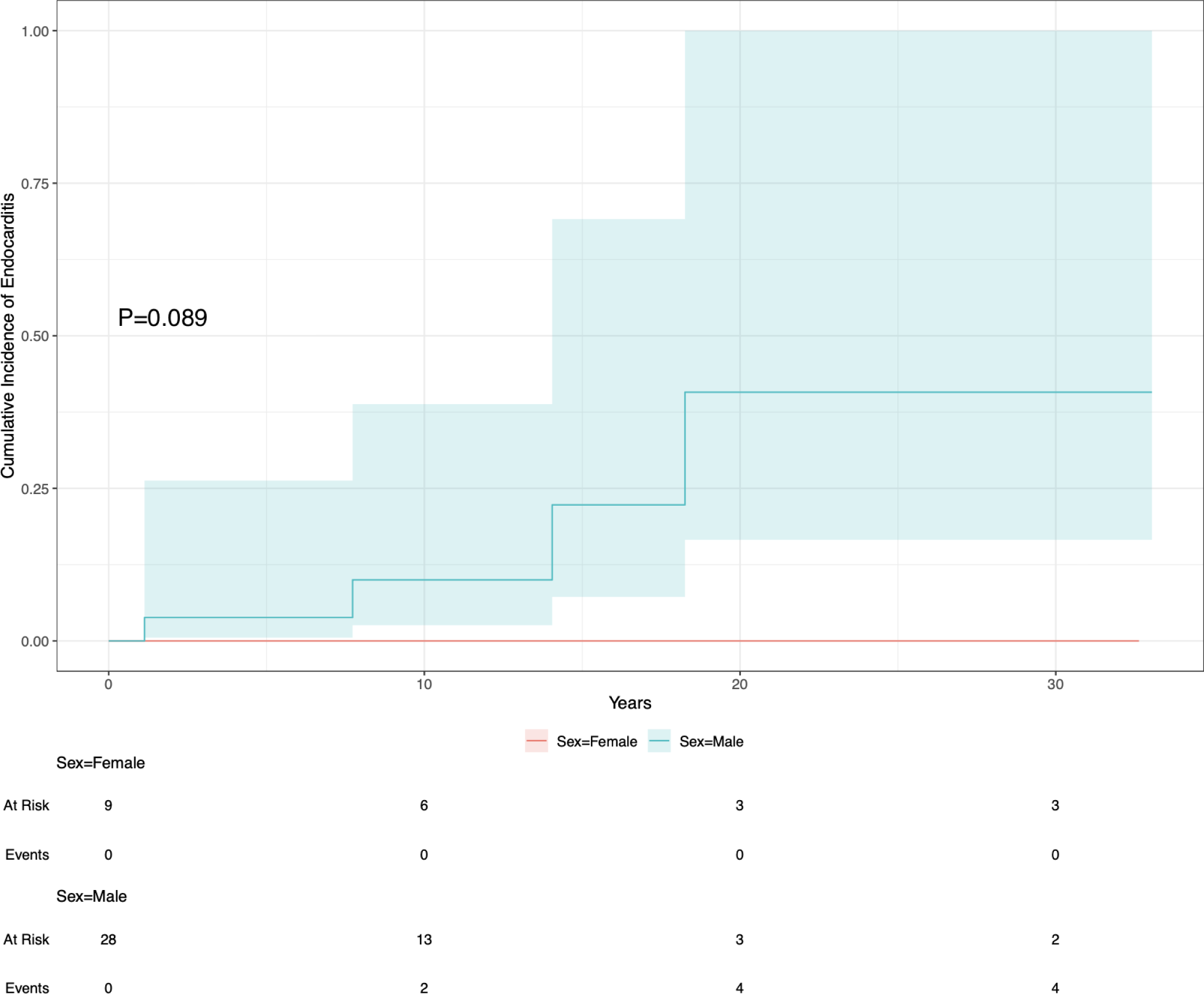
Cumulative incidence of homograft endocarditis in male and female patients.

### 3. Homograft function

The study encompassed 257 echocardiographs conducted on 33 homografts implanted in 20 patients(14 males, 6 females). The total echocardiographic follow-up time was 359.41 patient-years, with male patients contributing 264.14 patient-years and female patients contributing 95.27 patient-years. The completeness of echocardiographic follow-up was 83.2 %, calculated by the method proposed by Wu *et al*.^11^.Three patients moved abroad after the operations and no further echocardiographic information is available.

#### 3.1 Echocardiographic measurements

The evolution of RVOT peak gradient for each included patient was visualized and is presented respectively for male and female patients in Supplementary material Figure S6-S7. Different homografts in the same patient were plotted in the same panel using different lines.

Results of linear mixed-effects models for RVOT peak gradient are shown in Supplementary material Table S3. Male and female patients exhibited different rates of progression to stenosis, with males progressing at a faster rate (p=0.0272). The results of sensitivity analysis are presented in Supplementary material Figure S8, revealing significant disparities between the results of the five leave-one-homograft-out analyses and those of the full model. Results of logistic mixed-effects models for moderate or higher homograft regurgitation are displayed in Supplementary material Table S4, and its sensitivity analysis results are shown in Supplementary material Figure S9. The effect plots of RVOT peak gradient and probability of moderate or higher homograft regurgitation are presented in **Figure 5**.

**Figure 5:**
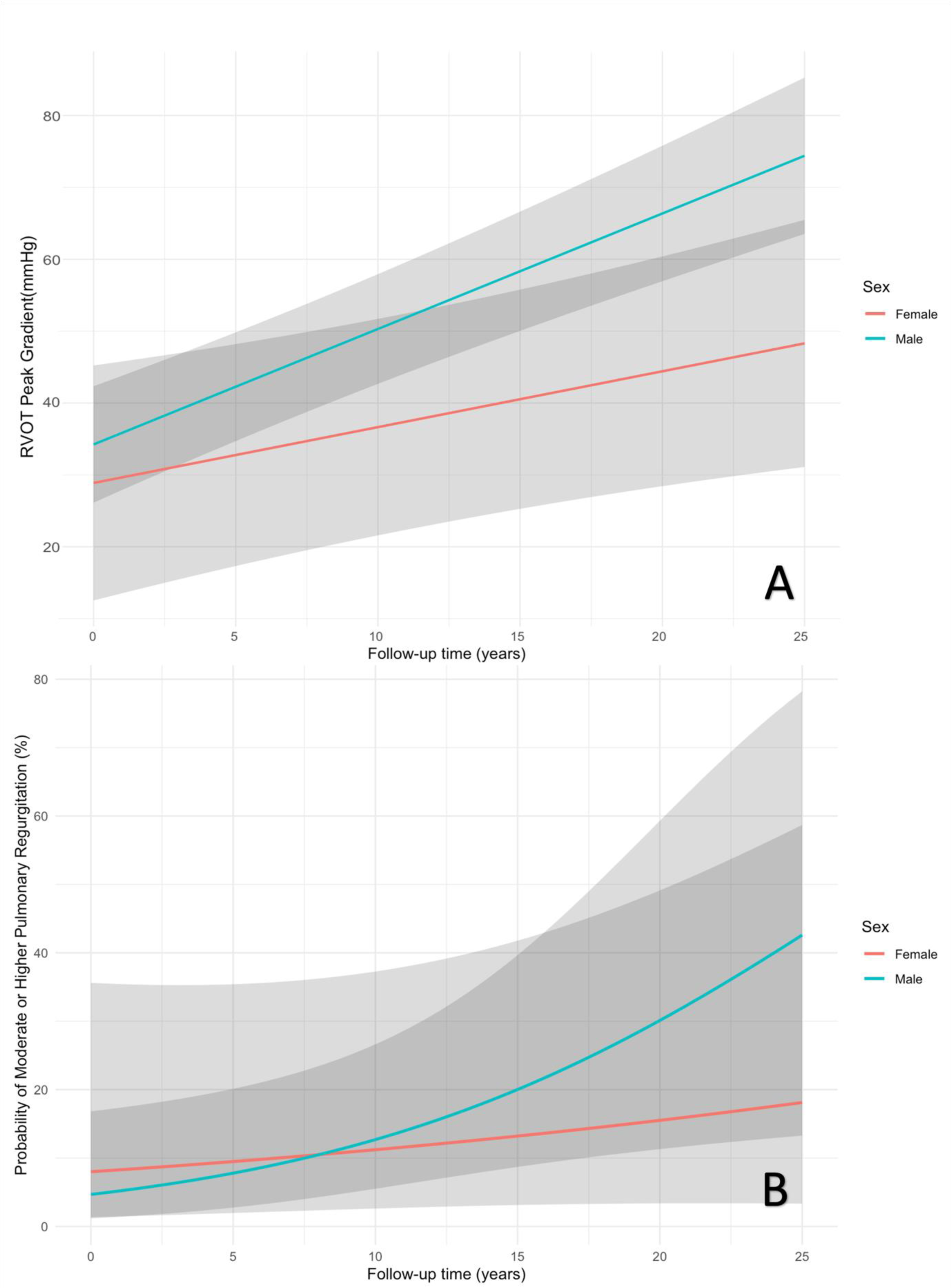
Effect plots of A: RVOT peak gradient change over time in male and female patients; B: probabilities of having moderate or higher pulmonary regurgitation over time in male and female patients.

#### 3.2 Associations between RVOT peak gradient and BSA in patients younger 16 years

One hundred eighty-seven BSA measurements were collected from 22 patients younger than 16 years when receiving homografts for RVOT reconstruction. Results of linear mixed-effects models for BSA evolution are presented in Supplementary material Table S5. The results of the two mixed-effects models for longitudinal evolutions of RVOT peak gradient and children’s BSA and their correlation matrix of random effects are exhibited in Supplementary material Table S6. The effect plot of BSA increase over time is shown in Supplementary material Figure S10. A faster increase of BSA over age was observed in boys compared to girls. The random slope of RVOT peak gradient over patient’s time-varying age was positively correlated to the slope of BSA over patient’s time-varying age (R = 0.5621). Furthermore, a linear regression between the increase of RVOT peak gradient and BSA over time revealed BSA growth rate was significantly association with RVOT peak gradient acceleration rate (7.27 [95% CI: 2.08-12.45], P = 0.0133). This association suggests the role of somatic growth in homograft stenotic failure. The effect estimate of BSA over time is presented for a 0.1 unite increase for its small scale.

## DISCUSSION

This study focused on male-female differences in clinical outcomes of d-TGA, VSD, PS/PA patients who received a homograft implantation for RVOT reconstruction, and the long-term course of homograft (dys)function. It shows that male patients have a higher rate of RVOT reintervention, especially reoperations, compared to female patients. Furthermore, a more rapid progression of homograft stenosis was observed in male patients, highlighting the significance of considering sex-specific factors when planning treatment and monitoring patient outcomes. On top of that, a strong association between the increase rates of BSA and RVOT peak gradient was found, underlining the necessity of accounting for the accelerated body growth experienced by young children. In light of this strong association, the use of oversized homografts during RVOT reconstruction might offer benefits for children with d-TGA, VSD, PS/PA, especially in younger children and boys, as they grow faster in body size.

Multiple factors may contribute to the observed male-female differences in this study, encompassing both biological (*sex*) factors such as body growth rate and hormonal influences, and socio-cultural (*gender*) aspects like societal norms and roles^12^. In order to comprehensively understand and utilize these differences in a clinically meaningful manner, it is essential to examine both sex and gender aspects within the context of clinical outcomes and homograft function.

### 1. Homograft reintervention

Many studies concentrating on outcomes of patients undergoing homograft replacement have found a higher rate of homograft reinterventions in males^13–15^, which is consistent with findings in this study. Several potential reasons may explain the phenomenon, e.g., anatomy and physiology, hormonal level, body size and growth rate, etc^16,17^.

Young children are more prone to early homograft degeneration compared to adults, due to prosthesis-patient mismatch developing over time, as a result of somatic growth^4,18^. In this study, most of the included patients received their first homograft for RVOT reconstructions before the age of 10, during which their body size rapidly increases over time. Consistent with previous research indicating an accelerated growth rate in young boys compared to girls^5^, our study cohort mirrored the same trend. The divergent growth rates may contribute to an increased occurrence of prosthesis-patient mismatch cases in male patients, and consequently, more reinterventions.

Furthermore, the observed male-female differences may be influenced by sex-specific hormones. During puberty, testosterone levels increase substantially in male adolescents, while estrogen levels rise dramatically in female adolescents^19,20^. Interestingly, these two sex steroid hormones have been found to be correlated with serum calcium levels^21^.Specifically, estrogen has been linked to reducing serum calcium, whereas testosterone has been associated with its elevation^21^. The potential impact of their regulatory roles on serum calcium could potentially impact the risk of homograft calcifications and degeneration.

On top of the aforementioned biological factors, gender may also contribute to the higher rate of reinterventions in male patients. Children’s gender might influence their parents’ care-seeking behavior for them, especially in low- and middle-income countries^22^. This may not directly apply to the current study, given that the Netherlands is a high-income country. Further information regarding patients and their families social-economic status would help validate whether children’s gender interacts with family income levels, potentially affecting parents’ healthcare-seeking decisions for their children.

Symptoms are an important part of criteria for heart valve reinterventions^7^. As indicated in some clinical research, male and female patients may differ in clinical symptoms, with female patients more likely to experience atypical symptoms^23–25^. The atypical symptoms might deter doctors from performing potentially necessary reinterventions in female patients, thereby causing a lower rate of reinterventions in female patients compared to male patients.

### 2. Homograft stenosis

In this study, out of the 22 reintervention cases, 18 were attributed to homograft stenotic failure. The RVOT peak gradient serves as a common echocardiographic parameter for assessing valve stenosis. Its evolution reflects the progression of stenosis. As depicted in **Figure 5**, male patients demonstrated a faster increase rate of RVOT peak gradient, implying a more rapid development of stenosis compared to their female counterparts. As previously discussed, prosthesis-patient mismatch could be a contributing factor to reinterventions for stenosis in young children. The observed faster stenotic development in male patients might be attributed to a higher incidence of prosthesis-patient mismatch in this group, as boys tend to grow faster than girls^5^. Our investigation of the relationship between the evolutions of BSA and RVOT peak gradient revealed a strong correlation, suggesting that faster BSA increase is correlated with a more rapid progression of RVOT peak gradient. Considering this strong correlation, accounting for children’s future body growth may become essential for clinicians, necessitating the consideration of different homograft sizing for boys and girls to provide more patient-specific clinical management.

Echocardiograph is commonly used to measure homograft function. Usually the time interval between two consecutive measurements depends on the decision made by both doctors and patients. As mentioned previously, female patients are inclined to exhibit more atypical symptoms or even remain asymptomatics^23–25^. On one hand, doctors may feel reluctant to prescribe appropriate treatment/examinations when facing patients with atypical symptoms. On the other hand, patients with non- or atypical-symptoms might be more likely to decline necessary homograft function evaluations, potentially leading to distortions of the observed evolutions of RVOT peak gradient. Consequently, (symptomatic) male patients received more echocardiographic examinations, capturing higher RVOT peak gradient values, which might contribute to the observed accelerated progression of homograft stenosis in male patients compared to their female counterparts.

### 3. Limitations

First and foremost, a notable limitation of this study is the very small sample size, which may have decreased the statistical power to detect the significant differences of the observation of many important clinical events during the follow-up period. Additionally, the application of complex modeling methods, such as (multivariate) mixed-effects models, in such a small dataset may lead to overfitted estimates, further warranting cautious extrapolation of results to other patients. Additionally, only linear relations between length of follow-up time and repeated measurements (RVOT peak gradient, ≥ moderate or higher pulmonary regurgitation) were taken into account to avoid potential overfitting issue. However, nonlinear relations between time and longitudinal outcomes are common in clinical research^26^. It is necessary to account for nonlinearity when larger sample size is available. Furthermore, reinterventions are recurrent events and should be analyzed using recurrent event survival analysis. However, it is not feasible due to the small sample size, and only the first reintervention was considered for survival analysis in this study. It might affect the estimates of reintervention hazard. Lastly, this is a retrospective study, and its inherent limitations also apply for this study^27^.

## Conclusion

In our single-center experience, there are substantial male-female differences in terms of clinical outcomes and homograft function over time in d-TGA, VSD, PS or PA patients who receive homografts for RVOT reconstruction during or after corrective operations. Given the association between the rate of body growth, notably faster in boys, and the progression of homograft stenosis, somatic growth might contribute to the higher reintervention rate in males. To address this, we recommend employing oversized homografts that account for future increases of body size in young patients when their body size can be measured accurately and the (properly) over-sized homograft is available. Furthermore, the oversizing criteria should be established sex-specifically in light of different body growth rates in young female and male patients.

## Data Availability

The data that support the findings of this study are available on request from the corresponding author, JJMT. The data are not publicly available due to the containing information that could compromise the privacy of research participants.

## ACKNOWLEDGMENTS

Not applicable

## AUTHOR CONTRIBUTION STATEMENT

**XW**: Conceptualization; Data collection and validation; Statistical analyses; Writing and editing. **IMB**: Conceptualization; Data collection and validation; Statistical analyses; Writing and editing. **KMV**: Methodological supervision; Results validation; Manuscript review and editing. **PCW**: Data validation; Manuscript review and editing. **CCEMV**: Data collection; Manuscript review and editing. **AJJCB**: Results validation; Manuscript review and editing. **WAH:** Results validation; Manuscript review and editing. **JJMT**: Conceptualization; Supervision; Data and results validation; Manuscript review and editing.

## REFERENCES

1. Carole AW. Transposition of the Great Arteries. Circulation. 2006;114(24):2699–2709. doi:doi:10.1161/CIRCULATIONAHA.105.592352

2. Hornung TS, Derrick GP, Deanfield JE, Redington AN. Transposition complexes in the adult: a changing perspective. Cardiology Clinics. 2002/08/01/ 2002;20(3):405–420. 10.1016/S0733-8651(02)00012-7

3. Romeo JLR, Mokhles MM, van de Woestijne P, et al. Long-term clinical outcome and echocardiographic function of homografts in the right ventricular outflow tract†. Eur J Cardiothorac Surg. Mar 1 2019;55(3):518–526.

4. Henaine R, Roubertie F, Vergnat M, Ninet J. Valve replacement in children: a challenge for a whole life. Arch Cardiovasc Dis. Oct 2012;105(10):517–28.

5. Kozieł S. Relationships among tempo of maturation, midparent height, and growth in height of adolescent boys and girls. Am J Hum Biol. Jan-Feb 2001;13(1):15–22.

6. Akins CW, Miller DC, Turina MI, et al. Guidelines for reporting mortality and morbidity after cardiac valve interventions. Ann Thorac Surg. Apr 2008;85(4):1490–5.

7. Vahanian A, Beyersdorf F, Praz F, et al. 2021 ESC/EACTS Guidelines for the management of valvular heart disease. Eur Heart J. Feb 12 2022;43(7):561–632.

8. Kim HY. Statistical notes for clinical researchers: Chi-squared test and Fisher’s exact test. Restor Dent Endod. May 2017;42(2):152–155.

9. Austin PC, Fine JP. Practical recommendations for reporting Fine-Gray model analyses for competing risk data. Stat Med. Nov 30 2017;36(27):4391–4400.

10. Rizopoulos D. The R Package JMbayes for Fitting Joint Models for Longitudinal and Time-to-Event Data Using MCMC. Journal of Statistical Software. 2016;72(7):1–46. doi:10.18637/jss.v072.i07

11. Wu Y, Takkenberg JJ, Grunkemeier GL. Measuring follow-up completeness. Ann Thorac Surg. Apr 2008;85(4):1155–7.

12. Bewley S, McCartney M, Meads C, Rogers A. Sex, gender, and medical data. Bmj. 2021;372:n735. doi:10.1136/bmj.n735

13. Oeser C, Uyanik-Uenal K, Kocher A, Laufer G, Andreas M. Long-term performance of pulmonary homografts after the Ross procedure: experience up to 25 years. Eur J Cardiothorac Surg. May 1 2019;55(5):876–884.

14. Caldarone CA, McCrindle BW, Van Arsdell GS, et al. Independent factors associated with longevity of prosthetic pulmonary valves and valved conduits. J Thorac Cardiovasc Surg. Dec 2000;120(6):1022–30; discussion 1031.

15. Zubairi R, Malik S, Jaquiss RDB, Imamura M, Gossett J, Morrow WR. Risk Factors for Prosthesis Failure in Pulmonary Valve Replacement. The Annals of Thoracic Surgery. 2011/02/01/ 2011;91(2):561–565. 10.1016/j.athoracsur.2010.07.111

16. Schoen FJ, Levy RJ. Calcification of tissue heart valve substitutes: progress toward understanding and prevention. Ann Thorac Surg. Mar 2005;79(3):1072–80.

17. Jiao L, Machuki JO, Wu Q, et al. Estrogen and calcium handling proteins: new discoveries and mechanisms in cardiovascular diseases. Am J Physiol Heart Circ Physiol. Apr 1 2020;318(4):H820–H829.

18. David TE. Aortic Valve Replacement in Children and Young Adults∗. Journal of the American College of Cardiology. 2016/06/21/ 2016;67(24):2871–2873. 10.1016/j.jacc.2016.04.023

19. Courant Fdr, Aksglaede L, Antignac J-P, et al. Assessment of Circulating Sex Steroid Levels in Prepubertal and Pubertal Boys and Girls by a Novel Ultrasensitive Gas Chromatography-Tandem Mass Spectrometry Method. The Journal of Clinical Endocrinology & Metabolism. 2010;95(1):82–92. doi:10.1210/jc.2009-1140

20. Khan L. Puberty: Onset and Progression. Pediatr Ann. Apr 1 2019;48(4):e141–e145.

21. Van Hemelrijck M, Michaelsson K, Nelson WG, et al. Association of serum calcium with serum sex steroid hormones in men in NHANES III. Aging Male. Dec 2013;16(4):151–8.

22. Sharif AI, Amy M, Sufang G, Alyssa S, Sheeba H, Paul R. Gender-related differences in care-seeking behaviour for newborns: a systematic review of the evidence in South Asia. BMJ Global Health. 2019;4(3):e001309. doi:10.1136/bmjgh-2018-001309

23. Meischke H, Yasui Y, Kuniyuki A, Bowen DJ, Andersen R, Urban N. How women label and respond to symptoms of acute myocardial infarction: responses to hypothetical symptom scenarios. Heart Lung. Jul-Aug 1999;28(4):261–9.

24. Dempsey SJ, Dracup K, Moser DK. Women’s decision to seek care for symptoms of acute myocardial infarction. Heart Lung. Nov-Dec 1995;24(6):444–56.

25. Davis LL, Mishel M, Moser DK, Esposito N, Lynn MR, Schwartz TA. Thoughts and behaviors of women with symptoms of acute coronary syndrome. Heart Lung. Nov-Dec 2013;42(6):428–35.

26. Wang X, Andrinopoulou ER, Veen KM, Bogers A, Takkenberg JJM. Statistical primer: an introduction to the application of linear mixed-effects models in cardiothoracic surgery outcomes research-a case study using homograft pulmonary valve replacement data. Eur J Cardiothorac Surg. Sep 2 2022;62(4)

27. Bellomo R, Warrillow SJ, Reade MC. Why we should be wary of single-center trials. Crit Care Med. Dec 2009;37(12):3114–9.

